# Systematic investigations of COVID-19 in 283 cancer patients

**DOI:** 10.1101/2020.04.28.20083246

**Authors:** Jie Wang, Qibin Song, Yuan Chen, Zhijie Wang, Qian Chu, Hongyun Gong, Shangli Cai, Xiaorong Dong, Bin Xu, Weidong Hu, Qun Wang, Linjun Li, Jiyuan Yang, Zhibin Xie, Zhiguo Luo, Jing Liu, Xiuli Luo, Jie Ren, Zhiguo Rao, Xinhua Xu, Dongfeng Pan, Zuowei Hu, Gang Feng, Chiding Hu, Liqiong Luo, Hongda Lu, Ruizhi Ran, Jun Jin, Yanhua Xu, Yong Yang, Zhihong Zhang, Li Kuang, Runkun Wang, Youhong Dong, Jianhai Sun, Wenbing Hu, Tienan Yi, Hanlin Wu, Mingyu Liu, Jiachen Xu, Jianchun Duan, Zhengyi Zhao, Guoqiang Wang, Yu Xu, Jie He

**Affiliations:** State Key Laboratory of Molecular Oncology, Department of Medical Oncology, National Cancer Center/National Clinical Research Center for Cancer/Cancer Hospital, Chinese Academy of Medical Sciences & Peking Union Medical College, Beijing 100021, China; Cancer Center, Renmin Hospital of Wuhan University, Wuhan, Hubei 430060, China; Department of Oncology, Tongji Hospital, Tongji Medical College, Huazhong University of Science and Technology, Wuhan, Hubei 430030, China; Burning Rock Biotech, Guangzhou 510300, China; Cancer Center, Union Hospital, Tongji Medical College, Huazhong University of Science and Technology, Wuhan, Hubei 430022, China; Department of thoracic Surgery, Zhongnan Hospital of Wuhan University, Wuhan, Hubei 430071, China; Department of Oncology, The Fifth Hospital of Wuhan, Wuhan, Hubei 430050, China; Department of Oncology, Hubei Provincial Hospital of Integrated Chinese and Western Medicine, Wuhan, Hubei 430071, China; Department of Oncology, First Affiliated Hospital of Yangtze University, Jingzhou, Hubei 434023, China; Department of Respiratory and Critical Care Medicine, Xiaogan Hospital Affiliated to Wuhan University of Science and Technology, Xiaogan, Hubei 432100, China; Department of Oncology, Taihe Hospital, Hubei University of Medicine, Shiyan, Hubei 442000, China; Department of Oncology, Huanggang Central Hospital, Huanggang, Hubei 438000, China; Department of Oncology, Hubei Provincial Hospital of TCM, Wuhan, Hubei 430000, China; Department of Medical Oncology, General Hospital of The Yangtze River Shipping, Wuhan, Hubei 430010, China; Department of Oncology, General Hospital of Central Theater Command, People’s Liberation Army, Wuhan, Hubei 430070, China; Department of Oncology, Yichang Central People’s Hospital, Yichang, Hubei 443003, China; Department of Oncology, Suizhou Hospital, HuBei University of Medicine, Suizhou, Hubei 441300, China; Department of Oncology, Wuhan No.1 Hospital, Wuhan, Hubei 430022, China; Department of Oncology, Wuhan Fourth Hospital (Puai Hospital), Tongji Medical College, Huazhong University of Science and Technology, Wuhan, Hubei 430000, China; Department of Oncology, Affiliated Hospital of Jianghan University, Wuhan, Hubei 430015, China; Department of Oncology, Tianyou Hospital Affiliated to Wuhan University of Science and Technology, Wuhan, Hubei 430064, China; Department of Oncology, The Central Hospital of Wuhan, Tongji Medical College, Huazhong University of Science and Technology, Wuhan, Hubei 430014, China; Department of Oncology, The Central Hospital of Enshi Tujia and Miao Autonomous Prefecture, Enshi, Hubei 445000, China; Department of Oncology, Ezhou Central Hospital, Ezhou, Hubei 436000, China; Department of Oncology, Jingzhou Central Hospital, Jingzhou, Hubei 434020, China; Department of oncology, The Second Hospital of WISCO (Wuhan Iron and Steel Corporation), Wuhan, Hubei 430085, China; Department of Oncology, Gong’an County People’s Hospital, Jingzhou, Hubei 434300, China; Department of Oncology, Affliated Dongfeng Hospital, Hubei University of Medicine, Shiyan, Hubei 442008, China; Department of oncology, The first people’s hospital of Guangshui, Suizhou, Hubei 432700, China; Department of oncology, Xiangyang No.1 People’s Hospital, Hubei University of Medicine, Xiangyang, Hubei 441000, China; Department of Oncology, Hubei No.3 People’s Hospital, Wuhan, Hubei 430033, China; Department of Oncology, Huangshi Central Hospital of EDong Healthcare, Huangshi, Hubei 435000, China; Department of Oncology, Xiangyang Central Hospital, Hubei Univeristy of Medicine, Xiangyang, Hubei 441021, China; Department of Oncology, The First People’s Hospital of Jingmen, Jingmen, Hubei 448000, China; The No. 9 hospital of Wuhan, Wuhan, Hubei 430081, China; The Medical Department, 3D Medicines, Inc., Shanghai 201114, China; State Key Laboratory of Molecular Oncology, Department of Thoracic Surgery, National Cancer Center/National Clinical Research Center for Cancer/Cancer Hospital, Chinese Academy of Medical Sciences & Peking Union Medical College, Beijing 100021, China

**Author notes:** Contributed equally (Jie Wang, Qibin Song, Yuan Chen, Zhijie Wang, Qian Chu, Hongyun Gong, Shangli Cai). **Correspondence to:** Jie He, M.D., State Key Laboratory of Molecular Oncology, Department of Thoracic Surgery, National Cancer Center/ National Clinical Research Center for Cancer/Cancer Hospital, Chinese Academy of Medical Sciences & Peking Union Medical College, Beijing, China, 17 Pan-jia-yuan South Lane, Chaoyang District, 100021, Beijing, China.; Jie Wang, M.D., Ph.D., State Key Laboratory of Molecular Oncology, Department of Medical Oncology, National Cancer Center/National Clinical Research Center for Cancer/Cancer Hospital, Chinese Academy of Medical Sciences & Peking Union Medical College, Beijing, China, 17 Pan-jia-yuan South Lane, Chaoyang District, 100021, Beijing, China.

## Abstract

**Background:** Cancer patients are considered to be highly susceptible to viral infections, however, the comprehensive features of COVID-19 in these patients remained largely unknown. The present study aimed to assess the clinical characteristics and outcomes of COVID-19 in a large cohort of cancer patients.

**Design, Setting, and Participants:** Data of consecutive cancer patients admitted to 33 designated hospitals for COVID-19 in Hubei province, China from December 17, 2019 to March 18, 2020 were retrospectively collected. The follow-up cutoff date was April 02, 2020. The clinical course and survival status of the cancer patients with COVID-19 were measured, and the potential risk factors of severe events and death were assessed through univariable and multivariable analyses.

**Results:** A total of 283 laboratory confirmed COVID-19 patients (50% male; median age, 63.0 years [IQR, 55.0 to 70.0]) with more than 20 cancer types were included. The overall mortality rate was 18% (50/283), and the median hospitalization stay for the survivors was 26 days. Amongst all, 76 (27%) were former cancer patients with curative resections for over five years without recurrence. The current cancer patients exhibited worse outcomes versus former cancer patients (overall survival, HR=2.45, 95%CI 1.10 to 5.44, log-rank *p*=0.02; mortality rate, 21% vs 9%). Of the 207 current cancer patients, 95 (46%) have received recent anti-tumor treatment, and the highest mortality rate was observed in the patients receiving recent chemotherapy (33%), followed by surgery (26%), other anti-tumor treatments (19%), and no anti-tumor treatment (15%). In addition, a higher mortality rate was observed in patients with lymphohematopoietic malignancies (LHM) (53%, 9/17), and all seven LHM patients with recent chemotherapy died. Multivariable analysis indicated that LHM (*p*=0.001) was one of the independent factors associating with critical illness or death.

**Conclusions:** This is the first systematic study comprehensively depicting COVID-19 in a large cancer cohort. Patients with tumors, especially LHM, may have poorer prognosis of COVID-19. Additional cares are warranted and non-emergency anti-tumor treatment should be cautiously used for these patients under the pandemic.

## Introduction

Since the publicly reported coronavirus disease 2019 (COVID-19) caused by severe acute respiratory syndrome coronavirus-2 (SARS-CoV-2) infection in December, 2019^1^ and the subsequent evidences of its human-to-human transmission property released in January 2020,^2^ there has been an extremely rapid spread of the novel coronavirus around the globe. The World Health Organization (WHO) has declared a global health emergency on January 30, 2020.^3^ The first systematic description of the clinical characteristics of COVID-19 in China was reported on February, 2020, showing a fatality rate of 1.4% among the 1,099 Chinese patients.^4^ Till April 02, 2020, the novel viral infection has caused 82,735 and 855,446 cumulative cases in China and worldwide, with 3,327 and 43,955 reported death, respectively.^5^

Cancer patients, as a special population who usually have a high demand for intensive care and are frequently systematically immunosuppressed, are considered to be more susceptible to the infection.^6^ Accordingly, a considerable amount of cancer patients may have been subjected to COVID-19 during the pandemic, which has attracted increasing attention from the clinicians.^6–9^ Recent studies with small cohorts (n = 12 to 28) of SARS-CoV-2 infected cancer patients have proposed the potentially higher susceptibility and poorer prognosis of COVID-19 in cancer patients compared to the overall population.^6,8,10^ However, the study population in these reports are too limited to provide conclusive evidences, and the characteristics of COVID-19 in cancer patients remained largely unknown. Herein, we carried out the present large retrospective study for the comprehensive investigation of the clinical characteristics, prognosis, survival status, and potential risk factors of severe events of COVID-19 in a large cohort of Chinese cancer patients.

## Methods

### Study Population, Setting and Data Collection

Consecutive cancer patients who were newly admitted to 33 designated hospitals for COVID-19 in Hubei province, China between December 17, 2019 and March 18, 2020 were included. The follow-up cutoff date was April 02, 2020. The patients were admitted to the hospitals for suspicious or confirmed SARS-CoV-2 infection irrespective of their on-admission symptom levels, and all patients eventually got laboratory confirmed diagnosis with COVID-19. The collection and tests of clinical specimens, diagnosis, treatment (anti-tumor and anti-infections), examinations, and severity level assessment (asymptomatic, mild, moderate, severe, and critical cases) of COVID-19 were performed following the latest edition of Novel Coronavirus Pneumonia Diagnosis and Treatment Plan released by the National Health Commission & National Administration of Traditional Chinese Medicine by assessment. Nasopharyngeal swabs were collected for pathogenic test using RT-PCR assay to confirm the presence of 2019-nCoV nucleic acid.

Data of the demographic and clinicopathological characteristics, infection symptoms, examination results (laboratory tests, chest CT scan, etc.), clinical treatments, and survival outcomes were retrospectively collected from the electronic medical records and further reviewed and confirmed by two independent physicians from each hospital. The data were analyzed by two independent statisticians (GW and YuXu). All inconsistencies were reviewed and confirmed by the Medical Review Board composed of physicians from departments of oncology, thoracic surgery, and pulmonary and critical care medicine. Patient identity protection was maintained throughout the study.

### Study Definitions

Former cancer patients were defined as the patients who were diagnosed with malignancies and have received curative surgical resections over five years without recurrence till the hospital admission for COVID-19. Fever was defined as an axillary temperature of 37.5°C or higher. Acute respiratory distress syndrome (ARDS) was defined based on the interim guidance for the management of critical COVID-19 by WHO.^11^ Recent anti-tumor treatment were defined as the conventional anti-tumor treatment that were provided within 3 months before the hospital admission for COVID-19. The types of conventional antitumor treatment included: surgical resection, chemotherapy, radiotherapy, targeted therapy, immunotherapy, and combinational therapy, etc. Critical cases were defined as patients with ARDS and requiring mechanical ventilation or extracorporeal membrane oxygenation (ECMO), shock, acute heart failure, acute myocardial infarction, or acute renal failure. Hospitalization stay was defined as the date from hospital admission for COVID-19 till discharge. Remission was defined as either the decrease of at least one-level on disease severity, the change of nucleic acid test results from positive to negative, or the improved absorption of inflammation upon pulmonary imaging. The overall survival (OS) was defined as the time from the date of first clinical visit for suspicious infection of SARS-CoV-2 until death due to any cause.

### Statistical Analysis

The clinical pathological variables, symptoms, and chest CT scan results in the patients were statistically described. Continuous variables were described by median (IQR) and categorical variables were described by number (percentages). To assess the differences between different subset of population, Mann-Whitney test was performed for continuous variables and chi-square test was performed for categorical variables. For hospitalization stay, Kaplan-Meier curves were plotted and compared by using a log-rank test with hazard’s ratio determined by cox regression. Univariable and multivariable logistic regression were used to test the associations between different variables and the critical and death cases. For univariable analysis, baseline characteristics were tested with blood biochemistry variables transformed into categorical variables with the cutoffs upon the clinical relevance. Variables that achieved a significant level with *p* < 0.05 and an odds ratio (OR) > 3 in the univariable analysis were selected for multivariable logistic regression using stepwise forward conditional mode. When strong correlations were present among the variables that met the above criteria, only the one with the highest odds ratio entered the multivariable analysis. All reported *p* values were two-tailed, and *p* < 0.05 was considered statistically significant. The analyses were performed with the use of R software (version 3.5.0, R Foundation for Statistical Computing) and GraphPad Prism software (version 5.0, GraphPad Software Inc.).

## Results

### Clinical Characteristics

A total of 283 cancer patients who were admitted to 33 designated hospitals with laboratory confirmed diagnosis of COVID-19 were included in the study (**Table 1**). The median age of the overall patients was 63.0 (IQR, 55.0-70.0) years, and 50% were male patients. The overall population harbored more than 20 types of tumor, including 51 (18%) with lung cancer, 38 (13%) with breast cancer, and 34 (12%) with colorectal cancer, etc. The most common comorbidities were hypertension (33%), diabetes (14%), and cardiovascular disease (11%). At onset of illness, 6 (2%), 97 (34%), 108 (38%), and 72 (25%) of the patients were asymptomatic, with mild, moderate, and severe symptoms, respectively. Amongst all, 76 (27%) were former cancer patients who have undergone primary resection for over five years without relapse, 207 (73%) were current cancer patients, including 101 (49%) with locally advanced or advanced diseases. Compared with the former cancer patients, the current cancer patients were more likely to exhibit severe symptoms (29% vs 17%) on admission.

**Table 1.** Demographic, clinical, laboratory findings of cancer patients at the first clinical visit.

|  |  | **Total**<br>**(n = 283)** | **Former cancer**<br>**patients**<br>**(n = 76)** | **Current cancer**<br>**patients**<br>**(n = 207)** |
| --- | --- | --- | --- | --- |
| **Demographic and clinical characteristics** |  |  |  |  |
| Age |  | 63.0 (55.0-70.0) | 65.0 (60.0-73.0) | 62.0 (53.0-69.0) |
| Sex |  |  |  |  |
|  | Female | 142 (50) | 45 (59) | 97 (47) |
|  | Male | 141 (50) | 31 (41) | 110 (53) |
| Smoking history |  |  |  |  |
|  | Current | 11 (4) | 1 (1) | 10 (5) |
|  | Former | 54 (19) | 10 (13) | 44 (21) |
|  | Never | 216 (76) | 65 (86) | 151 (73) |
|  | NA | 2 (1) | 0 (0) | 2 (1) |
| Drinking history |  |  |  |  |
|  | Current | 11 (4) | 1 (1) | 10 (5) |
|  | Former | 41 (14) | 5 (7) | 36 (17) |
|  | Never | 210 (74) | 69 (91) | 141 (68) |
|  | NA | 21 (7) | 1 (1) | 20 (10) |
| Huanan market exposure |  |  |  |  |
|  | No | 254 (90) | 67 (88) | 187 (90) |
|  | Yes | 10 (4) | 4 (5) | 6 (3) |
|  | NA | 19 (7) | 5 (7) | 14 (7) |
| Symptoms at the first visit |  |  |  |  |
|  | Asymptomatic | 6 (2) | 1 (1) | 5 (2) |
|  | Mild | 97 (34) | 26 (34) | 71 (34) |
|  | Moderate | 108 (38) | 36 (47) | 72 (35) |
|  | Severe | 72 (25) | 13 (17) | 59 (29) |
| Pathology |  |  |  |  |
|  | Lung cancer | 51 (18) | 5 (7) | 46 (22) |
|  | Breast cancer | 38 (13) | 17 (22) | 21 (10) |
|  | Colorectal cancer | 34 (12) | 14 (18) | 20 (10) |
|  | Thyroid carcinoma | 23 (8) | 7 (9) | 16 (8) |
|  | Gastric cancer | 18 (6) | 5 (7) | 13 (6) |
|  | Cervical cancer | 11 (4) | 5 (7) | 6 (3) |
|  | Leukemia | 11 (4) | 2 (3) | 9 (4) |
|  | Urothelial cancer | 11 (4) | 4 (5) | 7 (3) |
|  | Lymphoma | 10 (4) | 2 (3) | 8 (4) |
|  | Esophageal cancer | 9 (3) | 0 (0) | 9 (4) |
|  | Hepatocellular carcinoma | 9 (3) | 1 (1) | 8 (4) |
|  | Head and neck cancer | 7 (2) | 4 (5) | 3 (1) |
|  | Ovarian cancer | 5 (2) | 1 (1) | 4 (2) |
|  | Prostate cancer | 5 (2) | 1 (1) | 4 (2) |
|  | Sarcoma | 5 (2) | 1 (1) | 4 (2) |
|  | Pancreatic cancer | 4 (1) | 0 (0) | 4 (2) |
|  | Renal cell carcinoma | 4 (1) | 0 (0) | 4 (2) |
|  | Others | 28 (10) | 7 (9) | 21 (10) |
| Recent anti-cancer treatment |  |  |  |  |
|  | No | 188 (66) | 76 (100) | 112 (54) |
|  | Surgery | 23 (8) | 0 (0) | 23 (11) |
|  | Chemotherapy | 46 (16) | 0 (0) | 46 (22) |
|  | Targeted therapy | 12 (4) | 0 (0) | 12 (6) |
|  | Others | 14 (5) | 0 (0) | 14 (7) |
| Diabetes | No | 244 (86) | 60 (79) | 184 (89) |
|  | Yes | 39 (14) | 16 (21) | 23 (11) |
| Hypertension | No | 189 (67) | 45 (59) | 144 (70) |
|  | Yes | 94 (33) | 31 (41) | 63 (30) |
| Cardiovascular comorbidity | No | 251 (89) | 64 (84) | 187 (90) |
|  | Yes | 32 (11) | 12 (16) | 20 (10) |
| Cerebrovascular comorbidity | No | 264 (93) | 66 (87) | 198 (96) |
|  | Yes | 19 (7) | 10 (13) | 9 (4) |
| COPD | No | 263 (93) | 75 (99) | 188 (91) |
|  | Yes | 20 (7) | 1 (1) | 19 (9) |
| Chronic liver disease | No | 269 (95) | 74 (97) | 195 (94) |
|  | Yes | 14 (5) | 2 (3) | 12 (6) |
| Chronic renal disease | No | 273 (96) | 72 (95) | 201 (97) |
|  | Yes | 10 (4) | 4 (5) | 6 (3) |
| **Laboratory findings** |  |  |  |  |
| Leukocyte | ×10<sup>9</sup>/mL | 5.46 (4.16-7.23) | 5.61 (4.53-6.84) | 5.37 (3.90-7.26) |
| Neutrophil | ×10<sup>9</sup>/mL | 3.86 (2.59-5.47) | 3.81 (2.77-4.84) | 3.86 (2.54-5.76) |
| Lymphocyte | ×10<sup>9</sup>/mL | 0.86 (0.57-1.34) | 0.98 (0.72-1.54) | 0.80 (0.52-1.28) |
| Monocyte | ×10<sup>9</sup>/mL | 0.42 (0.30-0.60) | 0.44 (0.37-0.61) | 0.39 (0.26-0.59) |
| Eosinophil | ×10<sup>9</sup>/mL | 0.03 (0.00-0.08) | 0.05 (0.01-0.09) | 0.02 (0.00-0.08) |
| Basophil | ×10<sup>9</sup>/mL | 0.01 (0.01-0.02) | 0.01 (0.01-0.02) | 0.01 (0.00-0.02) |
| Platelet | ×10<sup>9</sup>/mL | 188 (121-247) | 222 (169-272) | 179 (104-227) |
| ALT | U/L | 22.0 (13.0-36.8) | 22.3 (15.8-33.3) | 21.5 (12.0-38.4) |
| AST | U/L | 28.0 (19.0-39.0) | 25.0 (19.0-39.0) | 28.5 (19.0-39.0) |
| TBIL | μmol/L | 10.8 (7.88-14.8) | 11.1 (9.35-14.5) | 10.5 (7.40-15.0) |
| DBIL | μmol/L | 3.70 (2.50-5.43) | 3.40 (2.50-4.30) | 4.00 (2.60-6.20) |
| Urea | mM | 4.88 (3.62-6.90) | 5.00 (4.00-6.23) | 4.80 (3.40-7.47) |
| Creatinine | μM | 64.1 (53.0-84.0) | 64.5 (55.0-83.8) | 64.1 (52.0-84.0) |
| Prothrombin time | s | 12.8 (11.8-13.8) | 12.7 (11.6-13.4) | 12.8 (11.9-14.0) |
| APTT | s | 33.3 (28.7-38.4) | 32.9 (28.4-37.7) | 33.5 (28.8-38.6) |
| D-dimer | μg/mL | 0.83 (0.36-2.00) | 0.57 (0.24-1.54) | 0.93 (0.47-2.49) |
| CK-MB | U/L | 1.31 (0.60-6.20) | 1.41 (0.90-4.46) | 1.12 (0.50-8.00) |
| Troponin | ng/ml | 0.08 (0.01-3.20) | 0.02 (0.01-2.20) | 1.69 (0.01-3.60) |
| CRP | mg/L | 26.5 (5.82-70.2) | 15.8 (5.00-56.0) | 29.3 (6.20-72.4) |
| hsCRP | mg/L | 27.1 (5.00-64.3) | 22.2 (5.00-56.6) | 28.0 (5.00-68.0) |
| Procalcitonin | ng/mL | 0.11 (0.05-0.26) | 0.08 (0.03-0.13) | 0.12 (0.05-0.34) |

Data are median (IQR), or n (%). Abbreviations: NA=not applicable; COPD=chronic obstructive pulmonary disease; ALT=alanine aminotransferase; AST=aspartate aminotransferase; TBIL=total bilirubin. DBIL=direct bilirubin; APTT=activated partial thromboplastin time; CK-MB=creatine kinase-MB; CRP=C-reactive protein; hsCRP=high sensitivity C-reactive protein.

The most common symptoms on admission were fever (73%), cough (68%), fatigue (49%), sputum (41%), and dyspnea (40%) (supplementary table S1). Baseline chest CT scan presented ground glass opacity in 211 (75%) patients, interstitial thickening in 112 (40%) patients, consolidation in 71 (25%) patients, and reticular pattern in 60 (21%) patients, which was similarly observed in the former and current cancer patients (supplementary table S2).

### Prognosis and outcomes

For treatment against COVID-19, antiviral, antibacterial, hormone, and immunoglobulin therapies were provided for 262 (93%), 233 (82%), 113 (40%), and 86 (30%) patients, respectively. The clinical courses of the patients throughout hospitalization were provided for the overall population (figure 1), the former cancer patients (supplementary figure S1), and the current cancer patients (supplementary figure S2). The overall mortality rate was 18% (50/283), including 17% (47/283) deaths due to COVID-19. During hospitalization, 19 mild cases, 34 moderate cases, and 72 severe cases eventually developed severe (62 [22%]) and critical (63 [22%]) cases. Of the critical cases, 45 (71%) died by the last visit, accounting for 90% of the overall death cases (figure 1). The most common complications were ARDS (85, 30%), stroke (42, 15%), and acute heart failure (17, 6%). Non-invasive and invasive mechanical ventilations were provided for 51 (18%) and 18 (6%) patients, respectively. At the last follow up, there were 178 (63%) asymptomatic cases and 2 (1%) severe cases (figure 1).

**Figure 1.**
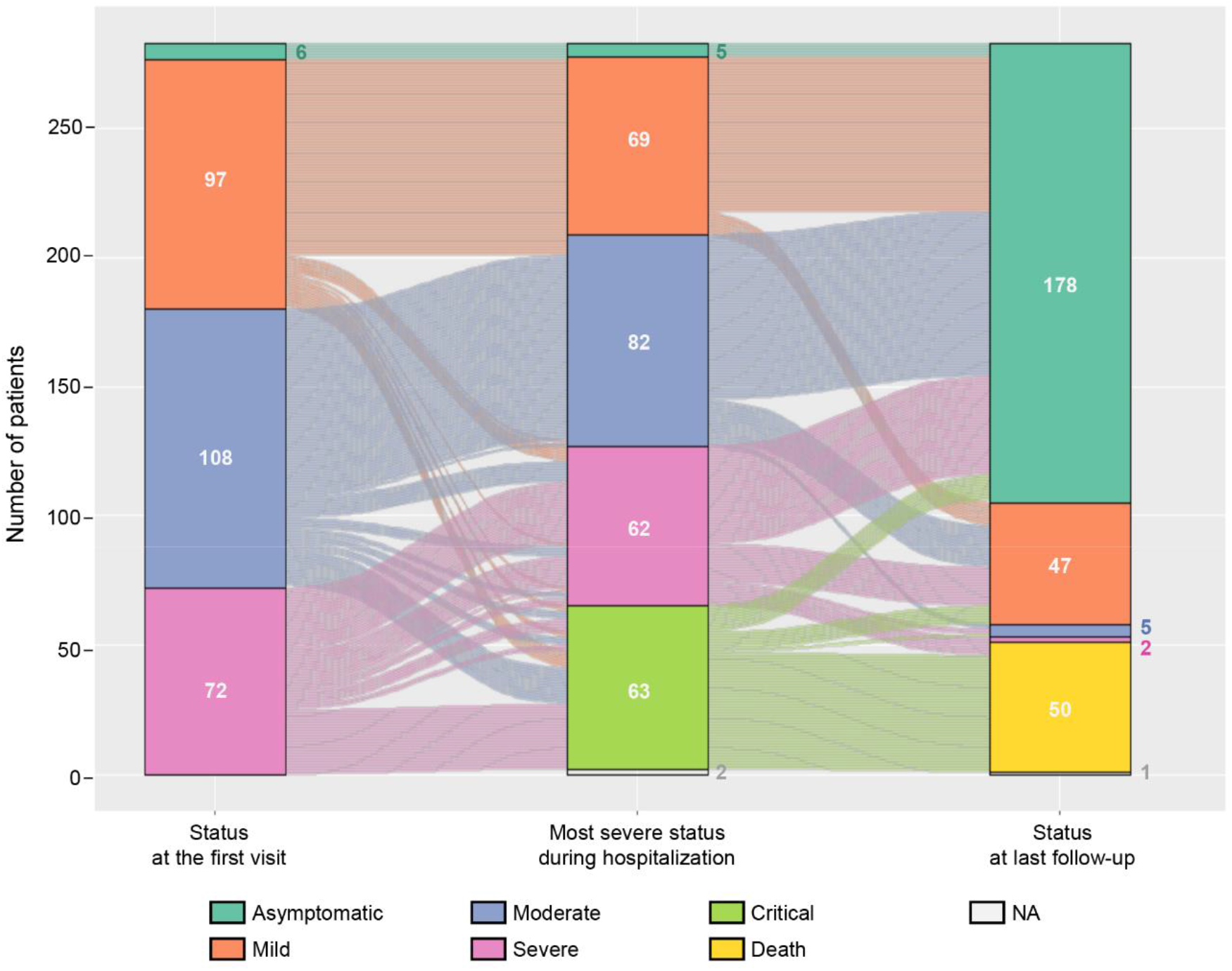
Clinical course of the patients in the study. Clinical evaluations of the patients were shown from their onset illness (left), most severe severity level (middle), to last follow-up visit (right). The patients with different severity levels were demonstrated in different colors.

The illustration of the clinical evaluations and outcomes of the patients with different types of cancer is provided in figure 2. The remission rate was 81% in the overall patients, 91% (69/76) in the former cancer patients, and 78% (161/207) in the current cancer patients, respectively. The recovery degree as represented by the ratio of asymptomatic patients by the last follow-up visit was 63% (178/283) in the overall population, 68% (52/76) in the former cancer patients, and 61% (126/207) in the current cancer patients, respectively. As shown, the worst prognosis was observed in the patients with current lymphohematopoietic malignancies (LHM) including lymphoma (n = 8) and leukemia (n = 9), followed by esophageal cancer (n = 9) and lung cancer (n = 51), with a remission rate of 38%, 44%, 67%, and 70%, and a degree of recovery of 38%, 22%, 44%, and 54%, respectively (figure 2, supplementary table S3). In addition, the LHM patients were more likely to develop severe or critical cases (59%, 10/17 vs 22%, 41/190) and deaths (53%, 9/17 vs 18%, 34/190) compared to patients with solid tumors (supplementary Table S4), and all the seven LHM patients with recent chemotherapy died.

**Figure 2.**
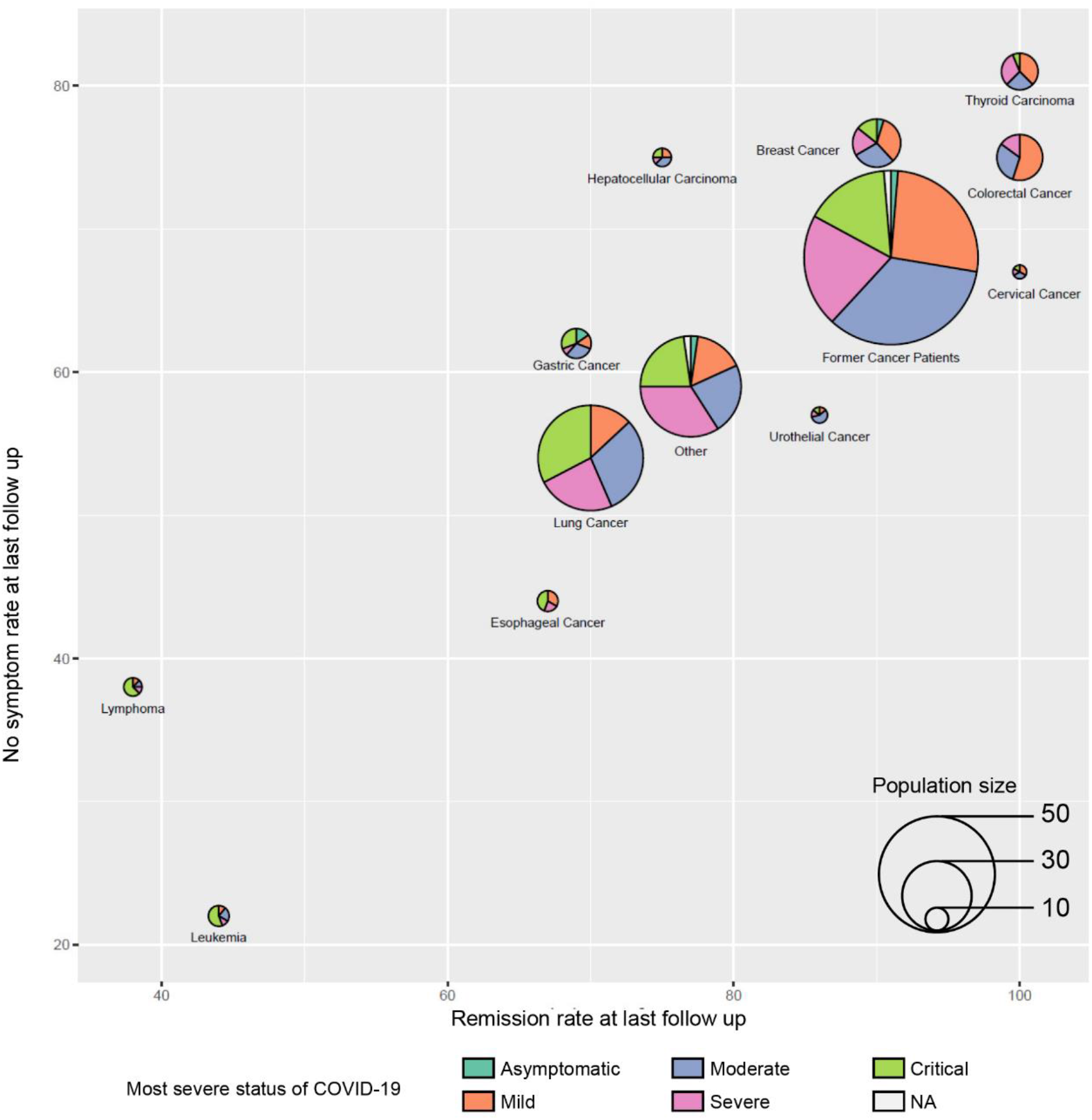
Illustration of the multidimensional outcomes of former cancer patients and the current cancer patients with different tumor types with COVID-19. Each pie represents one group of current cancer patient with a specific type of cancer, the former cancer patients were illustrated as a separate group. The y coordinate of each pie center represents the rate of patients with no symptoms at the last follow-up visit; the x coordinate represents the rate of remission at last follow-up visit; the composition within each pie represents the distribution of most severe symptom levels in each tumor type; the size of the pie represents the population size as demonstrated in the inset. Tumors with the population size less than 10 are not shown in the figure and were considered as “other”.

The median hospitalization stay was 24 days for the overall survivors (figure 3A) and the median duration from admission to death was 13.5 days in COVID-19 deceased cases (figure 3B). The current cancer patients exhibited a longer median hospitalization stay (26 days vs 25 days, figure 3C) and significantly higher risk of death compared to the former cancer patients (OS, HR 2.45, 95%CI 1.10-5.44, log-rank *p* = .023), with a mortality rate of 21% (43/207) and 9% (7/76), respectively (figure 3D). Of the 207 current cancer patients, 95 (46%) have received recent anti-tumor treatments, including 46 (22%) with chemotherapy, 23 (11%) with surgical resection, and 26 (13%) with other anti-tumor treatments. Among them, the highest mortality rate was observed in the patients treated with recent chemotherapy (33%), followed by surgery (26%), other treatments (19%), and no treatment (15%) (supplementary figure S3). Two patients have received recent immunotherapy as monotherapy, including one stage IV lung cancer patient with mild symptoms at diagnosis, experienced severe illness during hospitalization, got recovered after treatment and discharged with 31-day hospitalization stay, and another patient with stage IV urothelial cancer who had moderate symptoms as the worst status and got discharged after recovery with 26 days of hospitalization stay.

**Figure 3.**
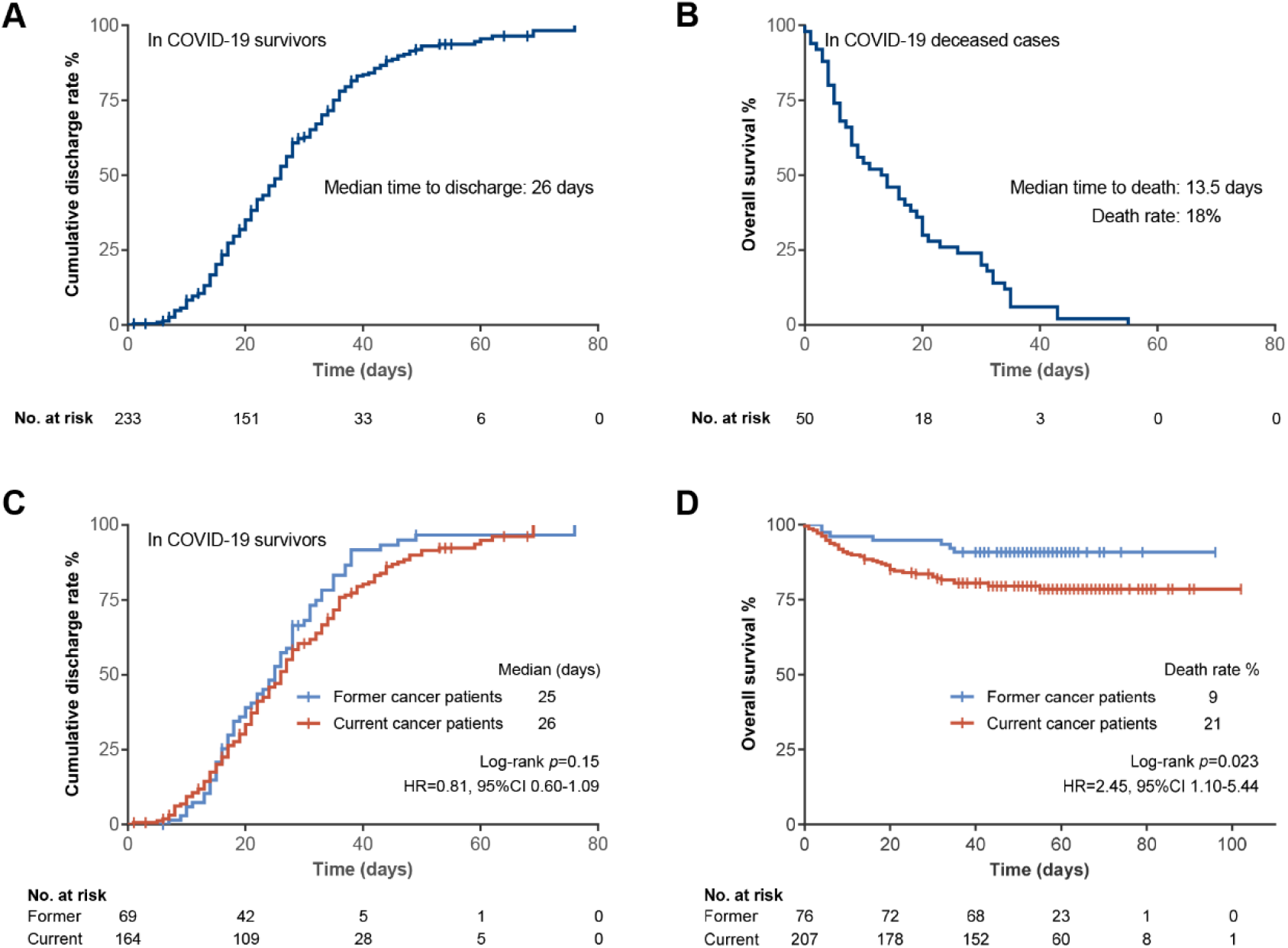
Kaplan-Meier curves of the outcomes of the patients. (A) Kaplan-Meier curves of the hospitalization stay of the overall survivors. (B) Kaplan-Meier curve of the time to death in COVID-19 deceased cases. (C) Kaplan-Meier curves comparing the hospitalization stay of survivors between the former and current cancer patients. (D) Comparison of the overall survival between the former and the current cancer patients.

### Risk factors

Logistic regression analyses were conducted to investigate the potential risk factors associated with disease severity in COVID-19 cancer patients. Demographic and clinical characteristics, symptoms, CT findings and laboratory findings were included for analysis. Multivariable analysis indicated that lymphohematopoietic malignancies (*p* = 0.001), severe symptoms at first clinical visit (*p* = 0.012), and elevated levels of neutrophil (> 6.3×10^9^/mL, *p* = 3.00×10^−6^), direct bilirubin (DBIL) (> 7 μmol/L, *p* = 0.001), creatinine (> 115 μM, *p* = 0.007), and troponin (> 5 ng/ml, *p* = 0.033) at baseline were independent risk factors associated with the development of critical illness or deaths (table 2). No significant association was observed between the cancer status (current vs former) with critical or death cases.

**Table 2.** Risk factors at the first clinical visit associated with critical ill or death.

|  |  |  | Univariable<br>OR (95%CI) | p value | Multivariable<br>OR (95%CI) | p value |
| --- | --- | --- | --- | --- | --- | --- |
| **Demographic and clinical characteristics** |  |  |  |  |  |  |
| Age | year | >65 vs. ≤65 | 1.94 (1.12-3.37) | 0.02 |  |  |
| Sex |  | Female vs. male | 0.67 (0.39-1.16) | 0.16 |  |  |
| Smoking history |  | No (ref.) |  |  |  |  |
|  |  | Current vs. ref. | 0.78 (0.16-3.72) | 0.75 |  |  |
|  |  | Former vs. ref. | 1.61 (0.83-3.10) | 0.16 |  |  |
| Drinking history |  | No (ref.) |  |  |  |  |
|  |  | Current vs. ref. | 0.75 (0.16-3.59) | 0.72 |  |  |
|  |  | Former vs. ref. | 1.40 (0.66-2.94) | 0.38 |  |  |
| Huanan market exposure |  | Yes vs. no | 0.77 (0.16-3.74) | 0.75 |  |  |
| Cancer history |  | Former (ref.) |  |  |  |  |
|  |  | Current vs. ref. | 1.98 (0.99-3.94) | 0.052 |  |  |
|  |  | LHM vs. ref. | 9.78 (3.03-31.51) | 1.34*10<sup>-4</sup> | 40.02 (4.35-368.69) | 0.001 |
|  |  | Lung cancer vs. ref. | 3.13 (1.32-7.38) | 0.009 | 1.69 (0.32-8.95) | 0.54 |
|  |  | Other cancers vs. ref. | 1.29 (0.61-2.70) | 0.51 | 1.00 (0.22-4.52) | 1.00 |
| Recent anti-cancer treatment |  | No treatment (ref.) |  |  |  |  |
|  |  | Any treatment vs. ref. | 1.93 (1.11-3.38) | 0.021 |  |  |
|  |  | Surgery vs. ref. | 1.43 (0.53-3.88) | 0.48 |  |  |
|  |  | Chemotherapy vs. ref. | 2.38 (1.18-4.78) | 0.015 |  |  |
|  |  | Others vs. ref. | 1.71 (0.69-4.20) | 0.24 |  |  |
| Diabetes |  | Yes vs. no | 0.42 (0.16-1.13) | 0.085 |  |  |
| Hypertension |  | Yes vs. no | 1.58 (0.90-2.78) | 0.11 |  |  |
| Cardiovascular comorbidity |  | Yes vs. no | 2.84 (1.33-6.07) | 0.007 |  |  |
| Cerebrovascular comorbidity |  | Yes vs. no | 0.83 (0.27-2.60) | 0.75 |  |  |
| COPD |  | Yes vs. no | 2.26 (0.88-5.77) | 0.090 |  |  |
| Chronic liver disease |  | Yes vs. no | 0.86 (0.23-3.16) | 0.82 |  |  |
| Chronic renal disease |  | Yes vs. no | 0.78 (0.16-3.78) | 0.76 |  |  |
| **Symptoms** |  |  |  |  |  |  |
| Symptoms at the first visit |  | Severe vs. non-severe | 5.10 (2.82-9.23) | 7.16*10<sup>-8</sup> | 5.43 (1.46-20.24) | 0.012 |
| Fever |  | Yes vs. no | 0.77 (0.42-1.42) | 0.40 |  |  |
| Cough |  | Yes vs. no | 1.51 (0.82-2.81) | 0.19 |  |  |
| Sputum |  | Yes vs. no | 2.12 (1.22-3.70) | 0.008 |  |  |
| Dyspnea |  | Yes vs. no | 4.14 (2.31-7.39) | 2.00*10<sup>-6</sup> |  |  |
| Fatigue |  | Yes vs. no | 1.74 (1.00-3.03) | 0.052 |  |  |
| Headache |  | Yes vs. no | 1.53 (0.56-4.19) | 0.41 |  |  |
| Muscle ache |  | Yes vs. no | 0.54 (0.20-1.46) | 0.22 |  |  |
| Sore throat |  | Yes vs. no | 1.18 (0.36-3.83) | 0.79 |  |  |
| Diarrhea |  | Yes vs. no | 0.63 (0.25-1.59) | 0.33 |  |  |
| Nausea |  | Yes vs. no | 0.46 (0.13-1.59) | 0.22 |  |  |
| Sneeze |  | Yes vs. no | 3.28 (0.45-23.72) | 0.24 |  |  |
| Nasal congestion |  | Yes vs. no | 1.07 (0.11-10.47) | 0.95 |  |  |
| Nasal discharge |  | Yes vs. no | 1.97 (0.46-8.46) | 0.36 |  |  |
| **CT findings** |  |  |  |  |  |  |
| Ground glass opacity |  | Yes vs. no | 0.72 (0.37-1.40) | 0.33 |  |  |
| Reticular pattern |  | Yes vs. no | 1.07 (0.55-2.08) | 0.85 |  |  |
| Interstitial thickening |  | Yes vs. no | 1.29 (0.73-2.27) | 0.38 |  |  |
| Crazy paving appearance |  | Yes vs. no | 1.06 (0.43-2.62) | 0.90 |  |  |
| Consolidation |  | Yes vs. no | 2.17 (1.19-3.96) | 0.011 |  |  |
| Bronchiectasis |  | Yes vs. no | 2.49 (1.05-5.91) | 0.039 |  |  |
| Bilateral involvement |  | Yes vs. no | 1.62 (0.71-3.69) | 0.25 |  |  |
| Multicentric pattern |  | Yes vs. no | 1.20 (0.64-2.25) | 0.58 |  |  |
| **Laboratory findings** |  |  |  |  |  |  |
| *Hematologic indices* |  |  |  |  |  |  |
| Leukocyte | ×10<sup>9</sup>/mL | >9.5 vs. ≤9.5 | 10.44 (4.75-22.95) | 5.23*10<sup>-9</sup> |  |  |
| Neutrophil | ×10<sup>9</sup>/mL | >6.3 vs. ≤6.3 | 9.56 (4.87-18.77) | 5.42*10<sup>-11</sup> | 32.77 (7.60-141.26) | 3.00*10<sup>-6</sup> |
| Lymphocyte | ×10<sup>9</sup>/mL | >1.1 vs. ≤1.1 | 0.34 (0.17-0.66) | 0.001 |  |  |
|  |  | >0.5 vs. ≤0.5 | 0.47 (0.25-0.92) | 0.03 |  |  |
| Monocyte | ×10<sup>9</sup>/mL | >0.6 vs. ≤0.6 | 2.95 (1.62-5.37) | 4.14*10<sup>-4</sup> |  |  |
| Eosinophil | ×10<sup>9</sup>/mL | >0.02 vs. ≤0.02 | 0.19 (0.10-0.36) | 0.00E0 |  |  |
| Basophil | ×10<sup>9</sup>/mL | >0.06 vs. ≤0.06 | 1.66 (0.48-5.72) | 0.42 |  |  |
| Platelet | ×10<sup>9</sup>/mL | >125 vs. ≤125 | 0.35 (0.19-0.62) | 3.94*10<sup>-4</sup> |  |  |
| *Hepatic function* |  |  |  |  |  |  |
| ALT | U/L | >40 vs. ≤40 | 1.47 (0.78-2.79) | 0.23 |  |  |
| AST | U/L | >40 vs. ≤40 | 2.62 (1.43-4.80) | 0.002 |  |  |
| TBIL | μmol/L | >17.1 vs. ≤17.1 | 4.27 (2.17-8.40) | 2.55*10<sup>-5</sup> |  |  |
| DBIL | μmol/L | >7 vs. ≤7 | 6.50 (3.25-13.01) | 1.20*10<sup>-7</sup> | 14.48 (2.79-75.22) | 0.001 |
| *Renal function* |  |  |  |  |  |  |
| Urea | mM | >7.1 vs. ≤7.1 | 5.24 (2.82-9.72) | 1.55*10<sup>-7</sup> |  |  |
| Creatinine | μM | >115 vs. ≤115 | 6.43 (2.75-15.03) | 1.72*10<sup>-5</sup> | 23.22 (2.40-224.35) | 0.007 |
| *Coagulation function* |  |  |  |  |  |  |
| Prothrombin time | s | >14 vs. ≤14 | 3.66 (1.92-6.98) | 8.17*10<sup>-5</sup> |  |  |
| APTT | s | >37 vs. ≤37 | 1.19 (0.65-2.18) | 0.56 |  |  |
| D-dimer | μg/mL | >0.55 vs. ≤0.55 | 3.40 (1.67-6.94) | 0.001 |  |  |
|  |  | >2 vs. ≤2 | 3.47 (1.86-6.47) | 8.80*10<sup>-5</sup> |  |  |
| *Myocardial injury* |  |  |  |  |  |  |
| CK-MB | U/L | >0.6 vs. ≤0.6 | 3.35 (1.33-8.43) | 0.010 |  |  |
|  |  | >6 vs. ≤6 | 2.86 (1.42-5.79) | 0.003 |  |  |
| Troponin | ng/ml | >0.1 vs. ≤0.1 | 1.30 (0.68-2.48) | 0.42 |  |  |
|  |  | >5 vs. ≤5 | 3.56 (1.64-7.71) | 0.001 | 5.57 (1.15-27.10) | 0.033 |
| *Infection-related indices* |  |  |  |  |  |  |
| CRP | mg/L | >10 vs. ≤10 | 5.60 (2.11-14.88) | 0.001 |  |  |
|  |  | >100 vs. ≤100 | 5.92 (2.70-13.01) | 9.00*10<sup>-6</sup> |  |  |
| hsCRP | mg/L | >5 vs. ≤5 | 3.38 (1.42-8.06) | 0.006 |  |  |
|  |  | >100 vs. ≤100 | 5.48 (2.25-13.38) | 1.87*10<sup>-4</sup> |  |  |
| Procalcitonin | ng/mL | >0.1 vs. ≤0.1 | 3.07 (1.60-5.91) | 0.001 |  |  |
|  |  | >0.5 vs. ≤0.5 | 5.56 (2.54-12.18) | 1.82*10<sup>-5</sup> |  |  |

*p* values were calculated by univariable or multivariable logistic regression. Abbreviations: NA=not applicable. COPD=chronic obstructive pulmonary disease. ALT=alanine aminotransferase; AST=aspartate aminotransferase; TBIL=total bilirubin; DBIL=direct bilirubin; APTT=activated partial thromboplastin time; CK-MB=creatine kinase-MB; CRP=C-reactive protein; hsCRP=high sensitivity C-reactive protein; LHM= Lymphohematopoietic cancer.

## Discussion

The rapid spreading of COVID-19 has swept the globe, leading to a worldwide pandemic. The present study has provided the first comprehensive depiction of the clinicopathological characteristics, clinical outcomes, and risk factors for death of COVID-19 in patients with tumors. Our study highlighted the poorer prognosis of COVID-19 in cancer patients, which may provide valuable clues for the patient management under the current coronavirus disease pandemic.

In our cohort of 283 cancer patients with COVID-19 infection, the median hospitalization stay was 26 days, which is longer than the previously reported 12 days^4^ in the unselected COVID-19 cases and 16 days in the severe cases.^12^ Recent study reported that the mortality rate was 28.6% in 28 SARS-CoV-2 infected cancer patients,^8^ our analysis further provided a mortality rate of 18%, with 9% in former cancer patients and 21% in current cancer patients, both higher than that of the overall cumulative mortality rate of COVID-19 cases as reported in Hubei province (4.7%) over the same period (as of April 02, 2020),^5^ indicating the worse outcomes in the patients with a history of cancer when infected by the novel coronavirus.

A statistically significantly shorter overall survival was observed in the current cancer patients compared to the former cancer patients, and the difference was larger when the patients received recent anti-tumor treatment, especially chemotherapy, which is most likely to weaken the immune system among different treatments. For patients with LHM, who often have a severely defective immune system upon the largely replacement of normal functional immune cells by malignant cells, a higher prevalence was observed in the studied cohort (6.0%, 3.2% for leukemia and 3.5% for lymphoma), which almost doubles that of the prevalence of LHM in the overall cancer population in China (3.8%, 1.8% for leukemia and 2.1% for lymphoma),^13^ indicating that these patients may be more susceptible to SARS-CoV-2 infection. In addition, an even higher rate of death cases was observed in patients with LHM (53%) compared with the patients with solid tumors (18%), and all of the LHM patients who were treated with recent chemotherapy died. Collectively, our results further suggested that the worse outcomes in SARS-CoV-2 infected cancer patients may be attributed to both the intrinsic immunocompromised status from the malignancies and the immunosuppression caused by active anti-tumor treatments. Accordingly, anti-tumor treatment, especially chemotherapy, should be used with great caution to reduce the potential negative effects on the patients.

Previous reports suggested that age and coexisting conditions were potential risk factors associated with disease severity.^14–16^ Given the higher median age in our cancer cohort (63 years) than the reported median age of 47-year-old in the general COVID-19 patients, the effect of age on the disease severity was weakened, providing a relatively lower level of significance (OR, 1.94, 95%CI 1.12-3.37) in the univariable analysis compared to the other variables (table 2). Our study further showed that LHM, an elevated baseline neutrophil, DBIL, creatinine, troponin, and the severe symptoms at onset illness were independently associated with the development of critical or death cases in cancer patients with COVID-19, which may provide important implications for the patient cares.

There are several limitations in this study. Firstly, this study was conducted in a retrospective setting. Nevertheless, the patient population in this study was relatively large, and the patients were consecutively included irrespective of their baseline characteristics including the clinical symptoms, which will reduce the potential bias of the study population. Secondly, this study focused on the investigation of patients with cancer infected with SARS-CoV-2, and no direct comparison was conducted between the infected cancer patients and the non-cancer patients. Despite that, comparisons have been made between the current and the former cancer patients, who were considered to exhibit relatively more likely to cancer-free patients as a reference. Besides, the studied population came from as many as 33 designated hospitals in Hubei province, China, which is considered to be fairly representative for the overall condition of COVID-19 cancer patients in Hubei province, and thus was compared with the data of general COVID-19 patients in Hubei province over the same period.

## Conclusion

Our study provided the first systematic investigations of the clinical characterization, outcomes, and risk factors for disease severity of COVID-19 patients with malignancies. The results indicated a potentially higher susceptibility and poorer prognosis of patients with a history of cancer when infected with SARS-CoV-2. Given that there are no confirmed effective drugs for the prevention or treatment of COVID-19 to date, strict preventions and special cares are warranted for patients with malignancies to minimize the potential risks and impacts from infections under the pandemic.

## Data Availability

The data that support the findings of this study are available from the corresponding author on reasonable request.

## Contributors

JW and JH contributed equally to this paper and are joint corresponding authors. JW, QS, YC, ZW, QC, HG, and SC are joint first authors. All corresponding and first authors contributed to study concept and design. JW, ZW, SC, QS, QC, GW, YuXu, and ZZhao contributed to the acquisition, analysis, and interpretation of data. JW, ZW, SC, GW, YuXu, and ZZhao wrote the first draft of the report. All authors approved the final version of the report. JH and JW were responsible for the integrity and accuracy of the data and is the guarantor. The corresponding authors attest that all listed authors meet authorship criteria and that no others meeting the criteria have been omitted.

## Ethical approval

This study was approved by the Ethics Committee of the National Cancer Center (No. 20/061–2257, Beijing, China). The requirement for informed patient consent was waived by the ethics committee due to the urgent of data collection.

## Data sharing

The manuscript’s guarantor affirms that the manuscript is an honest, accurate, and transparent account of the study being reported; that no important aspects of the study have been omitted; and that any discrepancies from the study as planned and registered have been explained.

## References

1. Huang C, Wang Y, Li X, et al. Clinical features of patients infected with 2019 novel coronavirus in Wuhan, China. Lancet. Feb 15 2020;395(10223):497–506.

2. Li Q, Guan X, Wu P, et al. Early Transmission Dynamics in Wuhan, China, of Novel Coronavirus-Infected Pneumonia. The New England journal of medicine. Mar 26 2020;382(13):1199–1207.

3. World Health Organization. Coronavirus diease (COVID-19) pandemic. 2020, 2020.

4. Guan WJ, Ni ZY, Hu Y, et al. Clinical Characteristics of Coronavirus Disease 2019 in China. The New England journal of medicine. Feb 28 2020.

5. Tencent Epidemic Situation Tracker. 2020, 2020.

6. Liang W, Guan W, Chen R, et al. Cancer patients in SARS-CoV-2 infection: a nationwide analysis in China. The Lancet. Oncology. Mar 2020;21(3):335–337.

7. Wang Z, Wang J, He J. Active and Effective Measures for the Care of Patients with Cancer During the COVID-19 Spread in China. JAMA oncology. 2020;In press.

8. Zhang L, Zhu F, Xie L, et al. Clinical characteristics of COVID-19-infected cancer patients: A retrospective case study in three hospitals within Wuhan, China. Annals of oncology: official journal of the European Society for Medical Oncology. Mar 26 2020.

9. Yu J, Ouyang W, Chua MLK, Xie C. SARS-CoV-2 Transmission in Patients With Cancer at a Tertiary Care Hospital in Wuhan, China. JAMA oncology. 2020.

10. Yu J, Ouyang W, Chua MLK, Xie C. SARS-CoV-2 Transmission in Patients With Cancer at a Tertiary Care Hospital in Wuhan, China. JAMA oncology. Mar 25 2020.

11. World Health Organization. Clinical management of severe acute respiratory infection when novel coronavirus (nCoV) infection is suspected. Interim guidance. 2020.

12. Cao B, Wang Y, Wen D, et al. A Trial of Lopinavir-Ritonavir in Adults Hospitalized with Severe Covid-19. The New England journal of medicine. Mar 18 2020.

13. Chen W, Zheng R, Baade PD, et al. Cancer statistics in China, 2015. CA: a cancer journal for clinicians. Mar-Apr 2016;66(2):115–132.

14. Chen N, Zhou M, Dong X, et al. Epidemiological and clinical characteristics of 99 cases of 2019 novel coronavirus pneumonia in Wuhan, China: a descriptive study. Lancet. Feb 15 2020;395(10223):507–513.

15. Bhatraju PK, Ghassemieh BJ, Nichols M, et al. Covid-19 in Critically Ill Patients in the Seattle Region - Case Series. The New England journal of medicine. Mar 30 2020.

16. Wu C, Chen X, Cai Y, et al. Risk Factors Associated With Acute Respiratory Distress Syndrome and Death in Patients With Coronavirus Disease 2019 Pneumonia in Wuhan, China. JAMA internal medicine. Mar 13 2020.

